# Association between the use of levodopa/carbidopa and the disease outcomes included in the National Alzheimer’s Coordinating Center Uniform Data Set

**DOI:** 10.1101/2024.12.04.24318183

**Authors:** Zsuzsa Sárkány, Joana Damásio, Sandra Macedo-Ribeiro, Pedro M. Martins

## Abstract

**INTRODUCTION:** This retrospective study investigates whether exposition to levodopa/carbidopa (LA/CA) medication is associated with modified Alzheimer’s disease (AD) trajectories.

**METHODS:** Multivariate analysis used cerebrospinal fluid (CSF) biomarker information included in the National Alzheimer’s Coordinating Center Uniform Data Set for subjects with normal cognition (NC), mild cognitive impairment (MCI) and dementia (DE). Survival analyses examined the progression to MCI/DE and death events.

**RESULTS:** LA/CA use is associated with lower levels of CSF amyloid beta, phosphorylated tau (P-tau) and total Tau. After adjusting for age, sex and APOE ε4 allele presence, that effect was quantified by negative coefficients of the fitted linear mixed models − P values <0.01 in all cases except for P-tau in the MCI subgroup (P=0.02). No similar effects were identified for other antiparkinsonian drugs. Exposition to LA/CA decreased the progression from MCI to DE (P=0.03).

**DISCUSSION:** The identified effects of LA/CA exposition on AD biomarkers and progression deserve further investigation in controlled clinical trials.

## 1. BACKGROUND

Levodopa/carbidopa (LA/CA) medication is used in the context of Parkinson’s disease (PD) to treat motor symptoms caused by dopamine deficiency. LA/CA can also be prescribed to Alzheimer’s disease (AD) patients who manifest Parkinsonian signs in addition to the behavioral and psychological symptoms characteristic of AD. A stronger link between the pathophysiology of AD and dopamine deficiency is, however, suggested in a network meta-analysis of data from a total of 512 AD patients and 500 healthy controls.^1^ Dopaminergic dysfunction is furthermore confirmed in TgF344 rat, 3xTg-AD mouse, Tg2576 mouse and 5xFAD mouse models of AD as a result of, for example, serotonin-receptor blockade, impairment in the dopamine D2/3 receptor signalling, and loss of dopaminergic neurons in the ventral tegmental area.^2–4^ Protecting the dopaminergic system emerges, therefore, as a possible hypothesis for AD therapy.^5^

Levodopa (L-3,4-dihydroxyphenylalanine, L-DOPA) is a dopamine precursor that, unlike dopamine, is able to cross the blood-brain barrier. Orally administered levodopa can be prematurely converted into peripheral dopamine by the aromatic L-amino acid decarboxylase (AADC) enzyme. Combining levodopa with the AADC inhibitor carbidopa greatly increases the amount of levodopa available to the brain and decreases the required dose of oral levodopa. Motor complications such as delayed on or wearing off phenomena and dyskinesias are common after long-term treatment with levodopa.^6,7^ Dopamine agonists can be used either as adjunctive therapy or as levodopa-sparing agents to mimic the action of dopamine in stimulating striatal post-synaptic receptors.^6^ Dopamine agonists such as pramipexole, ropinirole and rotigotine have replaced levodopa as the first-line treatment of restless leg syndrome,^8^ a neurological disorder characterized by dopamine dysregulation rather than dopamine deficiency.^9^

The National Alzheimer’s Coordinating Center (NACC) developed and maintains the Uniform Data Set (UDS) of clinical information that has been collected from Alzheimer’s Disease Centers in the US since 2005.^10^ Among several other records, the UDS includes follow-ups of the clinical diagnosis and prescribed medication, and, in some cases, imaging and cerebrospinal fluid (CSF) biomarker data and the genetic characterization of the apolipoprotein E (APOE) genotype. Using the NACC-UDS resource, relationships have been established between CSF biomarkers, neuropsychiatric symptoms and trajectories of depression/apathy,^11,12^ patients misdiagnosed with AD and their medication use,^13^ metformin use and the risk of severe dementia in AD patients with type 2 diabetes,^14^ and vitamin D supplementation and dementia incidence rates,^15^ among others.^10^

Here we analyzed the CSF biomarker data and clinical trajectory of NACC-UDS participants who were prescribed LA/CA. Our goal was to investigate whether AD outcomes were affected by exposition to LA/CA while accounting for important variables such as the baseline diagnosis of dementia, subject demographics, and the presence of APOE allele ε4. Subgroups of individuals with normal cognition (NC), mild cognitive impairment (MCI) and dementia (DE) were separately assessed for the CSF biomarkers amyloid beta (A*β*42) − whose levels are negatively correlated with amyloid load in the brain −^16^ and CSF tau (both total and phosphorylated) − whose levels are positively correlated with neuronal damage and neurodegeneration.^17^ A hypothetical disease-progression effect of LA/CA was further tested by analyzing the probabilities of survival to cognitive decline and death events calculated for subjects with and without a history of LA/CA use. We found that LA/CA exposition was associated with lowered levels of all CSF biomarkers for AD and delayed progression from MCI to DE.

## 2. METHODS

### 2.1 Participants

The participants in this study were those included in the subset of the (de-identified) NACC-UDS sample (March 2024 data freeze) who allowed the sharing of research data with commercial entities. The NACC program was developed to facilitate collaborative involvement among Alzheimer’s Disease Research Centers (ADCs) in the US. In 2005, ADCs began collecting longitudinal demographic, clinical, neuropsychological, and diagnostic data using version 1 of the data set, which was subsequently updated and expanded with the implementation of versions 2 (2008) and 3 (2015) of the UDS.^10,18^ The UDS Version 3 is nonproprietary (available upon a data request) and provides a standardized methodology for assessing cognition and clinical characteristics of patients with AD and other neurological diseases.^10^ Each ADC enrols its participants according to its protocols, e.g., through clinician referral, self-referral by participants or family members, and active recruitment in community organizations. This longitudinal protocol requires annual follow-up while the participant is able and willing to be involved, and comprises 8 data-collection forms that are completed by clinicians or clinical staff in each ADC (https://naccdata.org).

### 2.2 Definition of Cases and Controls

Our study is divided into a multivariate analysis of CSF biomarker data and a survival analysis of disease progression data. Cases for which the baseline characterization of prescribed medication occurred more than 2 years after the CSF test were excluded (Fig. 1). Results from at least one CSF test were available for 1,942 subjects comprising the clinical subgroups diagnosed at the baseline with NC (972), MCI (293) and DE (677). For each clinical subgroup, LA/CA cases and controls were defined according to the absence (controls) or existence (cases) of reported use of levodopa or carbidopa medication. This criterion comprises current or past prescriptions of clinical drugs whose names include the words ‘levodopa’ or ‘carbidopa’ in any of the 40 ‘DRUG’ fields of UDS form A4. In an additional multivariate analysis, the population of LA/CA controls, i.e., participants who do not have reported use of LA/CA, were further divided into those who did (cases) or did not (controls) take any of the following non-LA/CA antiparkinsonian drugs: ‘pramipexole’, ‘ropinirole’, ‘bromocriptine’, ‘pergolide’, ‘cabergoline’, ‘tolcapone’, ‘rotigotine’, ‘entacapone’, or ‘rasagiline’ (Fig. 1). Each subgroup was characterized in terms of number of cases of Parkinsonian symptoms reported as PD, other parkinsonian disorder, Parkinsonian signs, or Parkinsonian gait disorder. In the survival analysis of cognitive decline (Fig. 2), subjects diagnosed with NC (13,442) or MCI (6,906) at the baseline were divided into LA/CA cases and controls following the same criterion as in the multivariate analysis above. Then, nearest-neighbour 1:1 matching was performed to obtain equally-sized LA/CA-exposed and LA/CA-naive samples in each NC and MCI groups. Disease progression was also characterized in terms of the probability of death events in the NC and MCI groups, and in an additional group of subjects diagnosed with DE at the baseline.

**Figure 1.**
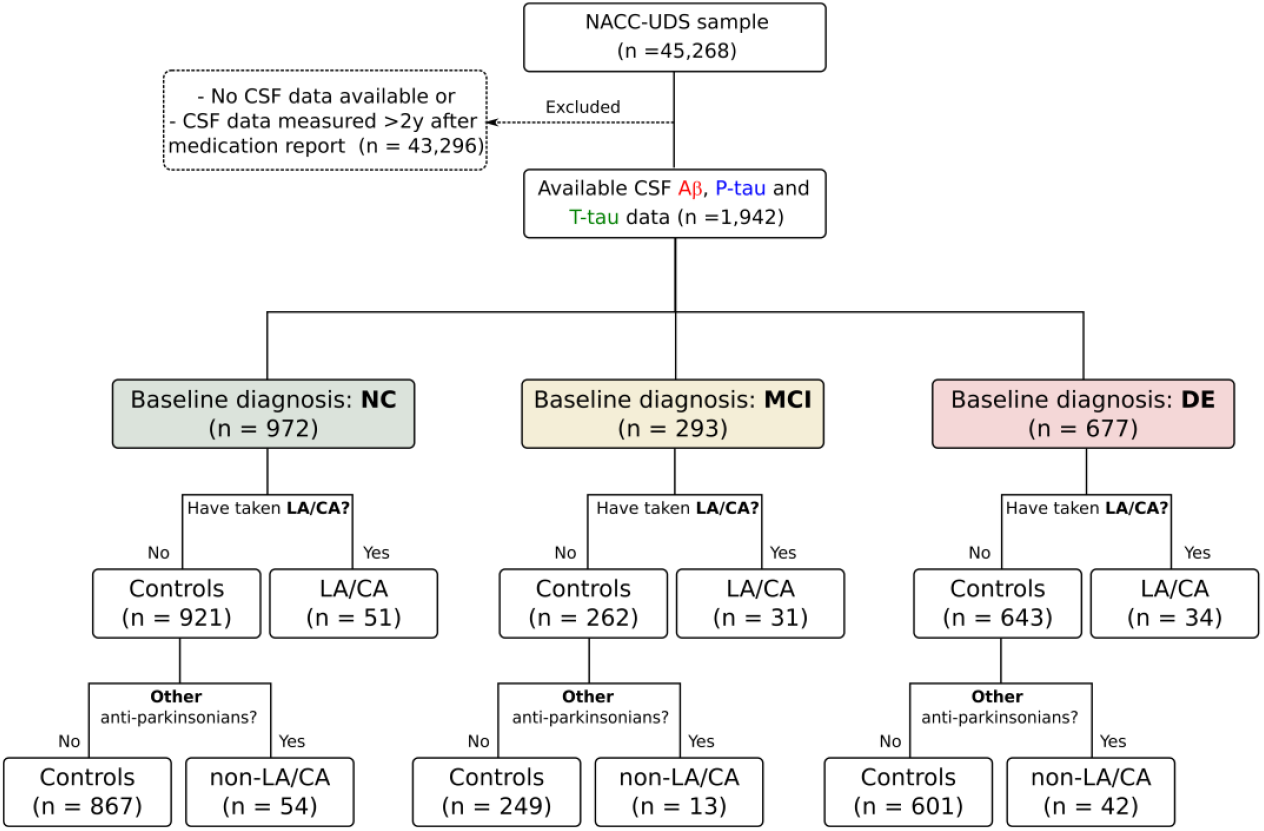
Flow chart of subject selection for multivariate analyses of CSF biomarker data in NC, MCI and DE subgroups exposed to LA/CA or other antiparkinsonian drugs, and controls.

**Figure 2.**
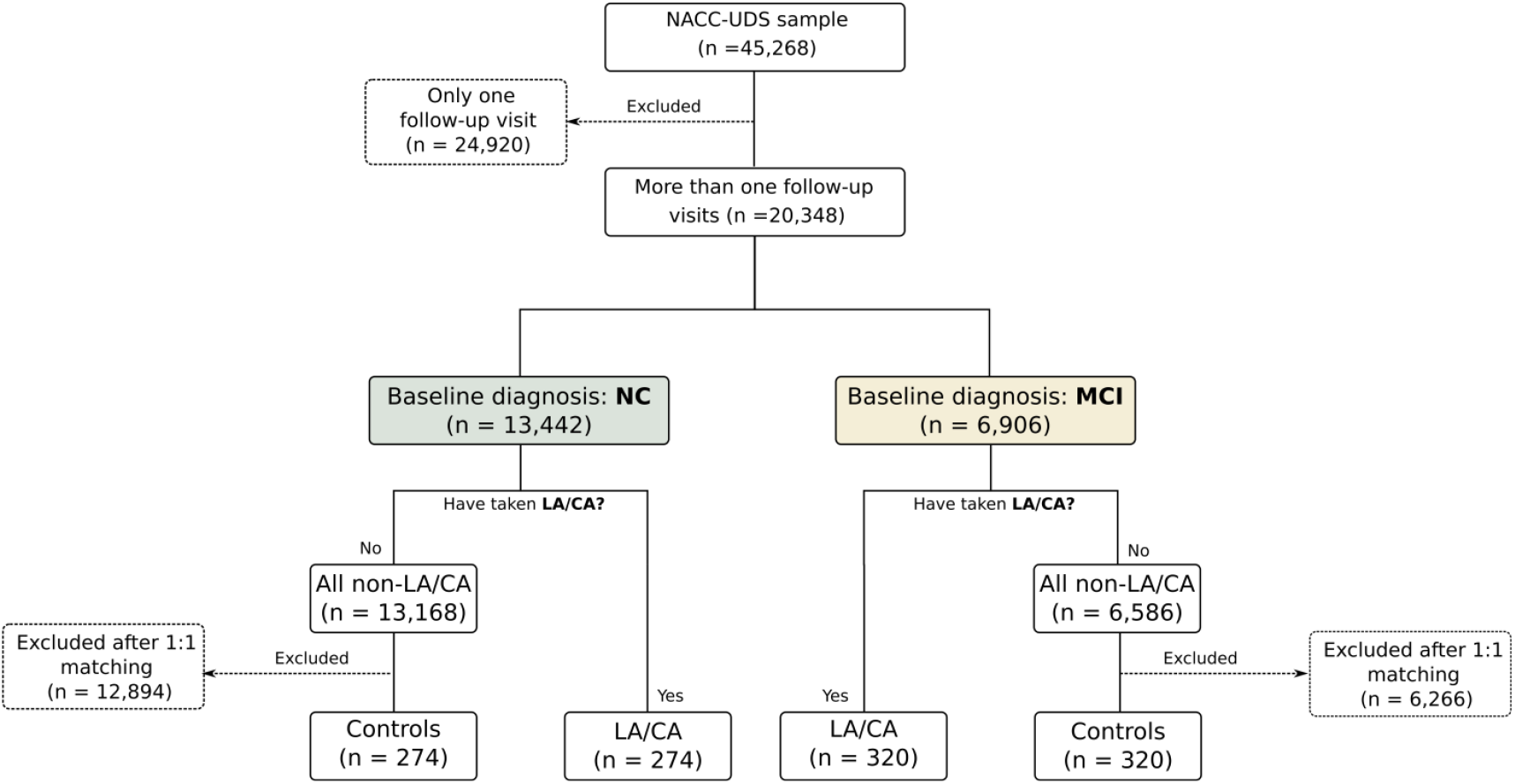
Flow chart of subject selection for the analyses of cognitve decline by NC and MCI subgroups exposed to LA/CA and controls.

### 2.3 Study Variables

The multivariate analysis uses CSF levels of A*β*42, total tau (T-tau), and tau phosphorylated at threonine 181 (P-tau) that are measured using enzyme-linked immunosorbent assay (ELISA) or Luminex multiplex xMAP assay protocols. Disease progression analyses used cognitive decline data obtained after clinical evaluation or the information of death events reported to NACC. The main covariates considered in our study were, for each clinical subgroup, age, sex and the presence of APOE allele ε4. Here, ‘APOE ε4 carriers’ classify the presence of APOE ε3/ε4, ε4/ε4 or ε4/ε2 genotypes. We also collected information about the education level (in years), race (grouped as White, Black or African American, and others), comorbidities such as hypertension, cardiovascular disease, and cancer, and concurrent medication such as nonsteroidal anti-inflammatory drugs (NSAIDs), anticoagulants, antipsychotics, hormones, antihypertensives, diabetes medication, lipid-lowering medication, and antidepressants.

### 2.4 Statistical Analysis

If not stated otherwise, the two-sided unpaired t-test or the Fisher test were carried out using the functions *ttest2* or *fishertest* of Matlab R2023a (Mathworks, Natick, MA) for comparisons between two groups of continuous variables or categorical variables, respectively. In our multivariate analysis, a linear mixed-effects (LME) model regression was performed to determine if the levels of CSF A*β*42, T-tau and P-tau are significantly affected by LA/CA use in the NC, MCI and DE subgroups. The adopted Wilkinson notation for the CSF levels of each biomarker was the following: BIOMARKER∼TREATMENT+AGE+SEX+APOE+(1|PATIENT), where TREATMENT denotes whether the patient was exposed to LA/CA or not, AGE, SEX, and APOE are additional covariates, and 1|PATIENT denotes random intercepts assigned to individual patients. The model was regressed using the *fitlme* function of MATLAB R2023a (MathWorks, Natick, MA) to determine LME coefficients (β) and the associated P values. In the survival analysis, the Kaplan-Meier method was used to determine the cumulative probabilities of (1) cognitive decline from NC to MCI/DE or from MCI to DE, and (2) the occurrence of death events in each clinical subgroup. Next, we performed propensity score matching and logrank (Mantel-Cox) tests to quantify the magnitude of the LA/CA effect. For that, the *NearestNeighbors*.*fit()* function from the Python package *scikit-learn* was first used for automatic 1:1 nearest-neighbour matching, and then GraphPad Prism software (La Jolla, CA, USA) was used to determine P values for each comparison of survival curves. P values less than 0.05 were considered significant.

## 3. RESULTS

### 3.1 Association between the use of LA/CA and CSF biomarker levels

LA/CA use was associated with lower levels of A*β*42, T-tau and P-tau in the CSF of subjects diagnosed at the baseline with NC, MCI or DE (Fig. 3, top). Although the populations of cases and controls were heterogeneously affected by different covariates (Table 1, top), the lowering effect of LA/CA on CSF biomarker levels was confirmed by LME model regressions adjusted for the effects of variables ‘age’, ‘sex’ and ‘presence of the APOE ε4 allele’ (Table 2, top). Statistically significant effects were also invariably observed for the APOE covariate in decreasing CSF A*β*42 levels and increasing CSF T-tau and P-tau. The effects of variable ‘age’ in increasing CSF T-tau and P-tau were statistically significant in all subgroups except DE. In our study, the variable ‘sex’ had no significant effect on biomarker levels except for CSF A*β*42 in the DE subgroup (CSF A*β*42 levels are lower in females diagnosed with DE). The percentages of subjects with reported Parkinsonian symptoms in the LA/CA cases/control groups were 27.4%/0.5% (NC), 32.2%/5.0% (MCI) and 14.7%/6.7% (DE).

**Table 1.** Characteristics of participants exposed to antiparkinsonian drugs (LA/CA, non-LA/CA and controls) in the CSF biomarker study.

|  | Variable | NC |  |  | MCI |  |  | DE |  |  |
| --- | --- | --- | --- | --- | --- | --- | --- | --- | --- | --- |
|  |  | Controls | Cases | P value | Controls | Cases | P value | Controls | Cases | P value |
| Levodopa/carbidopa (LA/CA) | n | 921 | 51 |  | 262 | 31 |  | 643 | 34 |  |
|  | Female n (%) | 517 (56.1) | 20 (39.2) | 0.021 | 108 (41.2) | 5 (16.1) | 0.006 | 293 (45.6) | 19 (55.9) | 0.29 |
|  | Age | 70 (9) | 67.3 (12) | 0.037 | 71.6 (9.1) | 70.4 (8.4) | 0.471 | 71.4 (9.4) | 71.6 (10.9) | 0.883 |
|  | ApoE4 n (%) | 338 (36.7) | 12 (23.5) | 0.071 | 122 (46.6) | 13 (41.9) | 0.705 | 336 (52.3) | 16 (47.1) | 0.6 |
|  | Education level (years) | 16.2 (4.7) | 15.9 (2.4) | 0.614 | 16 (6.2) | 16.3 (2.7) | 0.837 | 15.4 (7.4) | 14.6 (4.4) | 0.542 |
|  | Race/White n (%) | 822 (89.3) | 48 (94.1) | 0.352 | 236 (90.1) | 29 (93.5) | 0.751 | 581 (90.4) | 30 (88.2) | 0.564 |
|  | Race/Black or African American n (%) | 80 (8.7) | 1 (2) | 0.116 | 13 (5) | 1 (3.2) | 1 | 39 (6.1) | 1 (2.9) | 0.714 |
|  | Race/Other n (%) | 19 (2.1) | 2 (3.9) | 0.303 | 13 (5) | 1 (3.2) | 1 | 23 (3.6) | 3 (8.8) | 0.137 |
|  | Hypertension n (%) | 394 (42.8) | 15 (29.4) | 0.079 | 135 (51.5) | 14 (45.2) | 0.571 | 285 (44.3) | 15 (44.1) | 1 |
|  | Cardiovascular disease n (%) | 62 (6.7) | 9 (17.6) | 0.009 | 23 (8.8) | 3 (9.7) | 0.746 | 55 (8.6) | 5 (14.7) | 0.213 |
|  | Cancer n (%) | 32 (3.5) | 0 (0) | 0.406 | 11 (4.2) | 0 (0) | 0.613 | 19 (3) | 0 (0) | 0.617 |
|  | NSAIDs n (%) | 379 (41.2) | 7 (13.7) | 0 | 110 (42) | 7 (22.6) | 0.051 | 219 (34.1) | 6 (17.6) | 0.06 |
|  | Anticoagulant or antiplatelet agent n (%) | 379 (41.2) | 7 (13.7) | 0 | 110 (42) | 7 (22.6) | 0.051 | 219 (34.1) | 6 (17.6) | 0.06 |
|  | Antipsychotic agent n (%) | 11 (1.2) | 1 (2) | 0.478 | 12 (4.6) | 3 (9.7) | 0.203 | 36 (5.6) | 2 (5.9) | 1 |
|  | Hormone therapy n (%) | 44 (4.8) | 1 (2) | 0.508 | 8 (3.1) | 0 (0) | 1 | 13 (2) | 1 (2.9) | 0.517 |
|  | Antihypertensive or blood pressure n (%) | 427 (46.4) | 21 (41.2) | 0.564 | 148 (56.5) | 15 (48.4) | 0.446 | 317 (49.3) | 19 (55.9) | 0.486 |
|  | Diabetes medication n (%) | 65 (7.1) | 5 (9.8) | 0.406 | 38 (14.5) | 6 (19.4) | 0.434 | 54 (8.4) | 5 (14.7) | 0.207 |
|  | Lipid-lowering medication n (%) | 366 (39.7) | 15 (29.4) | 0.184 | 120 (45.8) | 14 (45.2) | 1 | 288 (44.8) | 14 (41.2) | 0.726 |
|  | Antidepressant n (%) | 221 (24) | 10 (19.6) | 0.612 | 103 (39.3) | 12 (38.7) | 1 | 265 (41.2) | 14 (41.2) | 1 |
|  | Variable | NC |  |  | MCI |  |  | DE |  |  |
|  |  | Controls | Cases | P value | Controls | Cases | P value | Controls | Cases | P value |
| Other Antiparkinsonians (non-LA/CA) | n | 867 | 54 |  | 249 | 13 |  | 601 | 42 |  |
|  | Female n (%) | 492 (56.7) | 25 (46.3) | 0.157 | 99 (39.8) | 9 (69.2) | 0.044 | 272 (45.3) | 21 (50) | 0.631 |
|  | Age | 70.1 (9) | 69.3 (7.9) | 0.537 | 71.7 (9) | 71.5 (12.4) | 0.945 | 71.4 (9.5) | 71.2 (8.3) | 0.883 |
|  | ApoE4 n (%) | 312 (36) | 26 (48.1) | 0.081 | 116 (46.6) | 6 (46.2) | 1 | 312 (51.9) | 24 (57.1) | 0.528 |
|  | Education level (years) | 16.2 (4.8) | 16.3 (2.8) | 0.878 | 15.7 (3.4) | 21.7 (23.4) | 0.001 | 15.2 (6.8) | 17.3 (13.4) | 0.078 |
|  | Race/White n (%) | 773 (89.2) | 49 (90.7) | 1 | 223 (89.6) | 13 (100) | 0.375 | 540 (89.9) | 41 (97.6) | 0.169 |
|  | Race/Black or African American n (%) | 77 (8.9) | 3 (5.6) | 0.616 | 13 (5.2) | 0 (0) | 1 | 39 (6.5) | 0 (0) | 0.101 |
|  | Race/Other n (%) | 17 (2) | 2 (3.7) | 0.307 | 13 (5.2) | 0 (0) | 1 | 22 (3.7) | 1 (2.4) | 1 |
|  | Hypertension n (%) | 371 (42.8) | 23 (42.6) | 1 | 132 (53) | 3 (23.1) | 0.046 | 270 (44.9) | 15 (35.7) | 0.265 |
|  | Cardiovascular disease n (%) | 60 (6.9) | 7 (13) | 0.104 | 23 (9.2) | 3 (23.1) | 0.127 | 52 (8.7) | 6 (14.3) | 0.257 |
|  | Cancer n (%) | 29 (3.3) | 3 (5.6) | 0.428 | 11 (4.4) | 0 (0) | 1 | 18 (3) | 1 (2.4) | 1 |
|  | NSAIDs n (%) | 362 (41.8) | 17 (31.5) | 0.155 | 104 (41.8) | 6 (46.2) | 0.78 | 203 (33.8) | 16 (38.1) | 0.614 |
|  | Anticoagulant or antiplatelet agent n (%) | 362 (41.8) | 17 (31.5) | 0.155 | 104 (41.8) | 6 (46.2) | 0.78 | 203 (33.8) | 16 (38.1) | 0.614 |
|  | Antipsychotic agent n (%) | 11 (1.3) | 0 (0) | 1 | 12 (4.8) | 0 (0) | 1 | 35 (5.8) | 1 (2.4) | 0.503 |
|  | Hormone therapy n (%) | 43 (5) | 1 (1.9) | 0.509 | 8 (3.2) | 0 (0) | 1 | 12 (2) | 1 (2.4) | 0.588 |
|  | Antihypertensive or blood pressure n (%) | 400 (46.1) | 27 (50) | 0.673 | 141 (56.6) | 7 (53.8) | 1 | 297 (49.4) | 20 (47.6) | 0.874 |
|  | Diabetes medication n (%) | 62 (7.2) | 3 (5.6) | 1 | 37 (14.9) | 1 (7.7) | 0.699 | 49 (8.2) | 5 (11.9) | 0.385 |
|  | Lipid-lowering medication n (%) | 342 (39.4) | 24 (44.4) | 0.477 | 113 (45.4) | 7 (53.8) | 0.58 | 267 (44.4) | 21 (50) | 0.523 |
|  | Antidepressant n (%) | 203 (23.4) | 18 (33.3) | 0.102 | 96 (38.6) | 7 (53.8) | 0.383 | 245 (40.8) | 20 (47.6) | 0.419 |

**Table 2.** Covariate-adjusted effect of (top) LA/CA and (bottom) non-LA/CA treatments on the AD biomarkers. LME coefficients, 95% confidence intervals and P values fitted to measured levels of A*β*42, T-tau and P-tau in the CSF of participants with NC, MCI and DE.

|  | Biomarker | EFFECT | NC |  |  |  | MCI |  |  |  | DE |  |  |  |
| --- | --- | --- | --- | --- | --- | --- | --- | --- | --- | --- | --- | --- | --- | --- |
| | | | $\beta$ | Lower | Upper | P value | $\beta$ | Lower | Upper | P value | $\beta$ | Lower | Upper | P value |
| Levodopa/carbidopa (LA/CA) | Aβ42 | INTERCEPT | 527.3 | 350.9 | 703.6 | <0.0001 | 188.8 | -73.5 | 451.0 | 0.1579 | 315.1 | 163.3 | 466.9 | 0.0001 |
|  |  | TREATMENT | -361.6 | -457.1 | -266.2 | <0.0001 | -136.1 | -236.7 | -35.5 | 0.0081 | -192.9 | -276.6 | -109.3 | <0.0001 |
|  |  | AGE | 1.2 | -1.0 | 3.5 | 0.2907 | 2.3 | -1.0 | 5.7 | 0.1723 | 1.8 | -0.1 | 3.8 | 0.0678 |
|  |  | SEX | 32.1 | -8.9 | 73.0 | 0.1245 | 48.1 | -14.2 | 110.5 | 0.1295 | -38.4 | -74.8 | -2.0 | 0.0388 |
|  |  | APOE4 | -93.2 | -114.5 | -71.9 | <0.0001 | -83.6 | -114.1 | -53.1 | <0.0001 | -61.9 | -80.8 | -42.9 | <0.0001 |
|  | P-tau | INTERCEPT | 9.5 | -5.2 | 24.2 | 0.2063 | 0.1 | -41.6 | 41.8 | 0.9968 | 71.1 | 40.5 | 101.6 | <0.0001 |
|  |  | TREATMENT | -15.8 | -23.7 | -7.9 | 0.0001 | -18.5 | -34.2 | -2.9 | 0.0204 | -30.7 | -47.5 | -14.0 | 0.0003 |
|  |  | AGE | 0.6 | 0.4 | 0.8 | <0.0001 | 0.7 | 0.2 | 1.3 | 0.0081 | -0.1 | -0.5 | 0.3 | 0.6596 |
|  |  | SEX | -1.9 | -5.3 | 1.6 | 0.2871 | 7.7 | -2.1 | 17.6 | 0.1248 | 4.1 | -3.2 | 11.5 | 0.2684 |
|  |  | APOE4 | 4.4 | 2.6 | 6.1 | <0.0001 | 14.9 | 10.1 | 19.8 | <0.0001 | 7.5 | 3.7 | 11.3 | 0.0001 |
|  | T-Tau | INTERCEPT | -51.5 | -178.8 | 75.9 | 0.4281 | -104.8 | -397.2 | 187.6 | 0.4816 | 457.4 | 206.6 | 708.2 | 0.0004 |
|  |  | TREATMENT | -204.5 | -273.5 | -135.5 | <0.0001 | -250.7 | -363.0 | -138.5 | <0.0001 | -316.4 | -454.6 | -178.2 | <0.0001 |
|  |  | AGE | 5.1 | 3.4 | 6.7 | <0.0001 | 6.5 | 2.7 | 10.2 | 0.0008 | -0.5 | -3.8 | 2.7 | 0.7372 |
|  |  | SEX | 0.7 | -28.8 | 30.3 | 0.9611 | -13.2 | -82.7 | 56.3 | 0.7083 | 8.8 | -51.4 | 68.9 | 0.7754 |
|  |  | APOE4 | 26.0 | 10.6 | 41.4 | 0.0009 | 66.9 | 32.9 | 100.9 | 0.0001 | 38.1 | 6.8 | 69.5 | 0.0171 |
|  | Biomarker | EFFECT | NC |  |  |  | MCI |  |  |  | DE |  |  |  |
| | | | $\beta$ | Lower | Upper | P value | $\beta$ | Lower | Upper | P value | $\beta$ | Lower | Upper | P value |
| Other Antiparkinsonians (non-LA/CA) | Aβ42 | INTERCEPT | 509.7 | 323.6 | 695.8 | <0.0001 | 6.4 | -9.3 | 22.1 | 0.4251 | -74.7 | -210.7 | 61.2 | 0.2812 |
|  |  | TREATMENT | -43.8 | -126.7 | 39.2 | 0.3011 | 1.5 | -5.5 | 8.5 | 0.6776 | -23.3 | -84.1 | 37.5 | 0.4521 |
|  |  | AGE | 1.6 | -0.8 | 4.0 | 0.1868 | 0.6 | 0.4 | 0.8 | <0.0001 | 5.4 | 3.7 | 7.2 | <0.0001 |
|  |  | SEX | 27.4 | -15.2 | 69.9 | 0.2070 | -1.8 | -5.4 | 1.8 | 0.3253 | 0.1 | -31.0 | 31.2 | 0.9951 |
|  |  | APOE4 | -95.9 | -117.9 | -73.8 | <0.0001 | 4.5 | 2.7 | 6.4 | <0.0001 | 27.2 | 11.0 | 43.3 | 0.0010 |
|  | P-tau | INTERCEPT | 113.4 | -166.6 | 393.3 | 0.4264 | -4.2 | -49.5 | 41.0 | 0.8540 | -142.0 | -460.1 | 176.1 | 0.3807 |
|  |  | TREATMENT | 145.1 | 8.8 | 281.4 | 0.0370 | -7.4 | -29.3 | 14.5 | 0.5062 | -125.7 | -280.5 | 29.1 | 0.1112 |
|  |  | AGE | 3.2 | -0.4 | 6.7 | 0.0850 | 0.8 | 0.2 | 1.4 | 0.0083 | 7.0 | 2.9 | 11.1 | 0.0008 |
|  |  | SEX | 53.8 | -12.4 | 120.1 | 0.1109 | 8.0 | -2.6 | 18.6 | 0.1391 | -9.9 | -85.2 | 65.3 | 0.7952 |
|  |  | APOE4 | -86.8 | -119.7 | -53.8 | <0.0001 | 16.2 | 10.9 | 21.5 | <0.0001 | 74.6 | 37.2 | 112.0 | 0.0001 |
|  | T-Tau | INTERCEPT | 290.1 | 130.6 | 449.6 | 0.0004 | 70.4 | 38.2 | 102.6 | <0.0001 | 460.1 | 196.5 | 723.8 | 0.0006 |
|  |  | TREATMENT | 69.6 | -1.5 | 140.7 | 0.0550 | -7.7 | -22.0 | 6.6 | 0.2921 | -93.7 | -211.2 | 23.8 | 0.1177 |
|  |  | AGE | 2.1 | 0.0 | 4.1 | 0.0493 | -0.1 | -0.5 | 0.4 | 0.7693 | -0.6 | -3.9 | 2.8 | 0.7488 |
|  |  | SEX | -36.8 | -74.9 | 1.3 | 0.0581 | 3.7 | -4.0 | 11.4 | 0.3490 | 11.8 | -51.1 | 74.8 | 0.7126 |
|  |  | APOE4 | -63.9 | -83.8 | -44.0 | <0.0001 | 7.9 | 3.9 | 11.9 | 0.0001 | 38.9 | 6.0 | 71.8 | 0.0204 |

**Figure 3.**
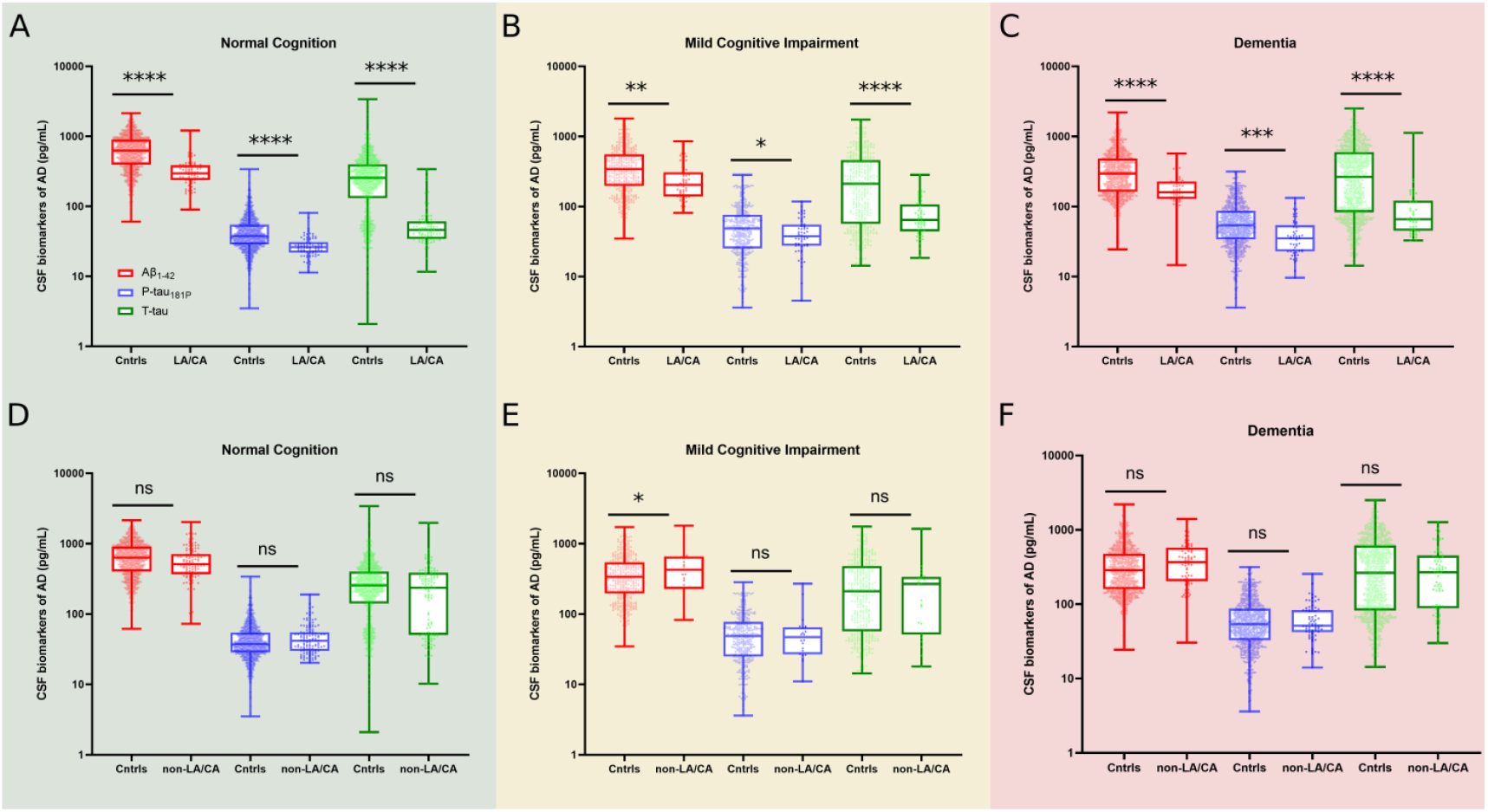
Association between the use of (top) LA/CA and (bottom) non-LA/CA antiparkinsonian drugs and the levels of CSF biomarkers for AD. Boxplots of measured levels of CSF A*β*42 (red), P-tau (blue) and T-tau (green) for the (A and D) NC, (B and E) MCI and DE (C and F) subgroups. The boxes extend from the 25th to 75th percentiles, the central line is the median and the whiskers represent minimum and maximum values. Stars show the statistical significance of differences between controls and cases quantified by LME model regression, with more stars indicating lower P values associated with the variable ‘TREATMENT’ in Table 2. Number of CSF tests: (A) 1481 controls and 79 LA/CA cases; (B) 377 controls and 54 LA/CA cases; (C) 886 controls and 53 LA/CA cases; (D) 1358 controls and 117 LA/CA cases; (E) 354 controls and 23 LA/CA cases; (E) 812 controls and 73 LA/CA cases.

We tested if the correspondence between lowered levels of AD biomarkers and LA/CA use is also observed in subjects prescribed dopamine agonists or other dopaminergic drugs distinct from LA/CA. None of the effects reported for LA/CA could be observed in the groups subject to levodopa-sparing treatments. No significant effect of non-LA/CA antiparkinsonian drugs was identified on the levels of CSF T-tau and P-tau in any of the subgroups NC, MCI or DE, while the CSF A*β*42 levels were increased (rather than decreased) in one of the subgroups (Fig. 3, bottom). As with the LA/CA results, a multivariate analysis adjusted for variables ‘age’, ‘sex’ and ‘presence of the APOE ε4 allele’ was performed to account for the heterogeneous populations of cases and controls (Table 1, bottom). Again, APOE ε4 carriers had systematically lower CSF A*β*42 and higher T-tau and P-tau levels, while older subjects of the subgroups NC and MCI tended to have higher T-tau and P-tau levels; no evident effect is identified for variable ‘sex’ (Table 2, bottom). The percentages of subjects with reported Parkinsonian symptoms in the non-LA/CA cases/control groups were 3.7%/4.0% (NC), 30.8%/3.6% (MCI) and 12.2%/6.0% (DE).

### 3.2 Association between the use of LA/CA and cognitive decline

Whereas the observation of lower CSF A*β*42 levels suggests changes in the A*β* metabolism, the T-tau- and P-tau-lowering effects might implicate reduced neurodegeneration and, consequently, slower cognitive decline. To test this hypothesis, all cases with reported use of LA/CA were assessed – including those without CSF biomarker data available – and a population of controls (without reported use of LA/CA) was defined upon automatic 1:1 matching by ‘age’, ‘sex’ and ‘APOE ε4 allele presence’ (Table 3). The Kaplan-Meier method was then used to determine the cumulative probabilities of cognitive decline from NC to MCI/DE (Fig. 4A) or from MCI to DE (Fig. 4B). According to the logrank Mantel-Cox tests that were performed, LA/CA exposition does not change the cognitive decline in NC subjects but increases the probability of no cognitive decline in MCI subjects (P=0.03; logrank hazard ratio 1.321 [95% confidence interval 1.020 to 1.710]).

**Table 3.** Characteristics of participants exposed to LA/CA and controls in the analyses of cognitive decline and survival rates.

|  | Variable | NC |  |  | MCI |  |  | DE |  |  |
| --- | --- | --- | --- | --- | --- | --- | --- | --- | --- | --- |
|  |  | Controls | Cases | P value | Controls | Cases | P value | Controls | Cases | P value |
| Levodopa/carbidopa (LA/CA) | n | 274 | 274 |  | 320 | 320 |  | 646 | 646 |  |
|  | Female n (%) | 132 (48.2) | 124 (45.3) | 0.549 | 81 (25.3) | 91 (28.4) | 0.422 | 195 (30.2) | 176 (27.2) | 0.268 |
|  | Age in years (SD) | 71.6 (8.9) | 70.5 (7.8) | 0.135 | 72.3 (9.9) | 70.8 (9.2) | 0.038 | 73.5 (9.6) | 72.2 (8.7) | 0.012 |
|  | Apoε4 n (%) | 58 (21.2) | 45 (16.4) | 0.189 | 64 (20) | 70 (21.9) | 0.627 | 208 (32.2) | 213 (33) | 0.812 |
|  | Education level in years (SD) | 17.2 (9) | 16.7 (5.6) | 0.433 | 15.7 (5.6) | 16.6 (8.6) | 0.124 | 15.6 (9) | 16.1 (7.4) | 0.327 |
|  | Race/White n (%) | 216 (78.8) | 258 (94.2) | <0.001 | 274 (85.6) | 300 (93.8) | 0.001 | 553 (85.6) | 597 (92.4) | <0.001 |
|  | Race/Black or African American n (%) | 36 (13.1) | 7 (2.6) | <0.001 | 27 (8.4) | 12 (3.8) | 0.020 | 37 (5.7) | 24 (3.7) | 0.115 |
|  | Race/Other n (%) | 22 (8) | 9 (3.3) | 0.025 | 19 (5.9) | 8 (2.5) | 0.047 | 56 (8.7) | 25 (3.9) | 0.001 |
|  | Hypertension n (%) | 128 (46.7) | 114 (41.6) | 0.263 | 168 (52.5) | 127 (39.7) | 0.001 | 338 (52.3) | 320 (49.5) | 0.344 |
|  | Cardiovascular disease n (%) | 48 (17.5) | 38 (13.9) | 0.290 | 51 (15.9) | 57 (17.8) | 0.598 | 110 (17) | 101 (15.6) | 0.547 |
|  | Cancer n (%) | 16 (5.8) | 7 (2.6) | 0.086 | 24 (7.5) | 12 (3.8) | 0.058 | 38 (5.9) | 14 (2.2) | 0.001 |
|  | NSAIDs n (%) | 102 (37.2) | 121 (44.2) | 0.117 | 140 (43.8) | 133 (41.6) | 0.632 | 210 (32.5) | 234 (36.2) | 0.178 |
|  | Anticoagulant or antiplatelet agent n (%) | 102 (37.2) | 121 (44.2) | 0.117 | 140 (43.8) | 133 (41.6) | 0.632 | 210 (32.5) | 234 (36.2) | 0.178 |
|  | Antipsychotic agent n (%) | 4 (1.5) | 7 (2.6) | 0.545 | 6 (1.9) | 9 (2.8) | 0.603 | 54 (8.4) | 68 (10.5) | 0.216 |
|  | Hormone therapy n (%) | 11 (4) | 10 (3.6) | 1 | 3 (0.9) | 8 (2.5) | 0.223 | 8 (1.2) | 13 (2) | 0.379 |
|  | Antihypertensive or blood pressure n (%) | 143 (52.2) | 156 (56.9) | 0.303 | 189 (59.1) | 166 (51.9) | 0.08 | 332 (51.4) | 328 (50.8) | 0.867 |
|  | Diabetes medication n (%) | 29 (10.6) | 28 (10.2) | 1 | 52 (16.3) | 26 (8.1) | 0.002 | 73 (11.3) | 46 (7.1) | 0.012 |
|  | Lipid-lowering medication n (%) | 129 (47.1) | 105 (38.3) | 0.047 | 151 (47.2) | 129 (40.3) | 0.094 | 292 (45.2) | 293 (45.4) | 1 |
|  | Antidepressant n (%) | 41 (15) | 100 (36.5) | <0.001 | 87 (27.2) | 137 (42.8) | <0.001 | 271 (42) | 285 (44.1) | 0.465 |

**Figure 4.**
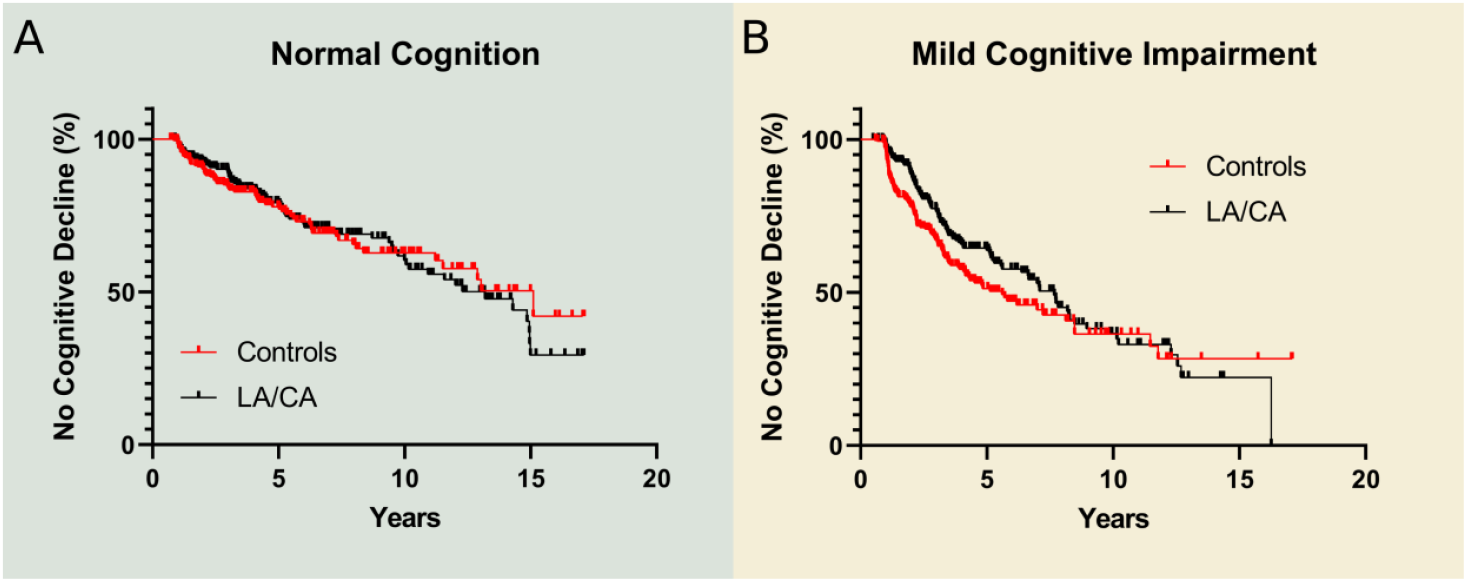
Resistance to cognitive decline by participants in the LA/CA-exposed and LA/CA-naive samples. Kaplan-Meier analysis for events of cognitive decline by subjects with (A) NC and (B) MCI.

### 3.2 Association between the use of LA/CA and registered death rates

We further speculated that the beneficial effects of LA/CA could also affect life expectancy. Kaplan-Meier curves were generated using the dates of deaths reported during the observation period (2005-2024) for the same NC and MCI populations that were assessed for cognitive decline. An analogous procedure was adopted for the subjects with baseline DE who were prescribed LA/CA (cases) and respective controls matched by ‘age’, ‘sex’ and ‘APOE ε4 allele presence’ (Table 3). In all these analyses, the probability of survival to death events was never increased in LA/CA-exposed groups (Fig. 5). Survival rates decreased in the NC subgroup (Fig. 5A, P=0.002; logrank hazard ratio 0.50 [95% confidence interval 0.334 to 0.763]) and remained unaltered in the MCI (Fig. 5B) and DE (Fig. 5C) subgroups.

**Figure 5.**
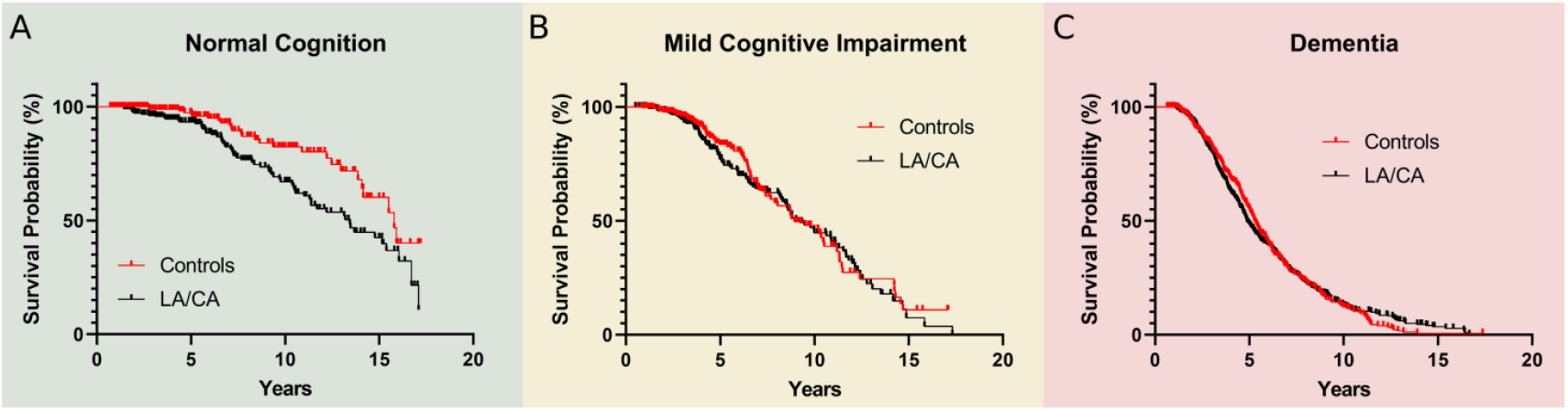
Overall survival of participants in the LA/CA-exposed and LA/CA-naive samples. Kaplan-Meier analysis for death events by subjects with (A) NC, (B) MCI and (C) DE.

## 4. DISCUSSION

Our study retrospectively compares CSF biomarker levels, cognitive decline, and probability of death events among LA/CA-exposed and LA/CA-naive groups at different stages of cognitive impairment due to AD. Lower levels of CSF A*β*42, T-tau- and P-tau were consistently observed in LA/CA-exposed subjects diagnosed at the baseline with NC, MCI or DE. This association, which was adjusted for important covariates such as the presence APOE ε4 allele or age, was not verified for levodopa-sparing antiparkinsonians such as dopamine agonists. Remarkably for MCI patients who show both PD and AD symptoms, LA/CA exposition was additionally associated with delayed progression to dementia. On the other hand, no effect of LA/CA use was observed on the probabilities of cognitive decline by NC subjects. The survival analysis of registered death events indicates no effect of LA/CA exposition in the MCI and DE subgroups and increased death rates in the NC subgroup. The survival analyses of cognitive decline and death events always use populations with the same number of cases and controls matched by the initial diagnostic of cognitive impairment (NC, MCI or DE), age, sex and presence APOE ε4 allele.

Previous works using animal models of AD offer plausible explanations for the possible effects of LA/CA on disease progression. Using a transgenic mouse model, Ambrée et al. showed that levodopa treatment ameliorates learning and memory deficits, increases the dopamine levels in the neostriata and frontal cortices, and decreases the dopamine levels in the hippocampi.^19^ Later on, Guzmán-Ramos et al. observed that memory impairment was also attenuated after restoration of dopamine release deficits through retrodialysis administration of nomifensine.^20^ It was argued that the increased availability of dopamine in specific regions of the brain may restore the dopaminergic equilibrium and the working memory impairment, and contribute to correcting the pronounced acetylcholine dysfunction characteristic of AD.^19,20^ Sub-chronic treatment with levodopa of transgenic mice showing a selective vulnerability of dopaminergic neurons in the ventral tegmental area (VTA) completely rescued CA1 synaptic plasticity and dendritic spine density, and restored hippocampal post-synaptic density composition, memory performance and food reward processing.^4^ Moreover, levodopa, as well as the dopamine D2 receptor agonists quinpirole and sumanirole, were able to ameliorate hippocampal hyperexcitability caused by degeneration of dopaminergic VTA neurons.^21^

A disease-modifying effect of LA/CA might also be linked with the main histopathological hallmarks of AD, viz. the extracellular accumulation of the amyloid-*β* peptide (A*β*) in amyloid plaques and the intracellular deposition of protein tau. It was recently shown *in vivo* that levodopa promotes the degradation of A*β* in a neprilysin-dependent manner,^22,23^ and improves cognitive function,^23^ but it does not affect tau pathology in 5xFAD mice.^22^ Lower levels of CSF A*β*42 in LA/CA-exposed subjects could alternatively be explained by an increased accumulation of amyloid plaques.^16^ Although A*β* aggregation is inhibited by dopamine,^24^ metabolites arising from dopamine oxidation can stabilize A*β* oligomers and thus remodel the amyloid cascade of events.^25^ Moreover, abnormal metabolism of A*β*42 may be a common feature of PD patients with MCI or DE.^26^ Our observations of reduced levels of A*β*42, T-tau and P-tau associated with exposition to LA/CA but not dopamine agonists shed some light on the potential use of CSF biomarkers in PD diagnosis. Although there is no consensus on how these biomarkers correlate with PD in studies not controlled for antiparkinsonian use,^26–30^ reduced levels of the A*β* peptide and tau proteins are confirmed in a cohort of entirely untreated PD patients compared to healthy controls.^28^ Furthermore, our multivariate analysis underlines the importance of the APOE ε4 genotypes in reducing A*β*42 levels and increasing T-tau and P-tau levels.

We showed that LA/CA treatments are associated with reduced levels of biomarkers for A*β*42 metabolism and tau pathology. This relationship is not observed for levodopa-sparing antiparkinsonians and goes along with a significant delay in the progression to dementia of MCI patients, especially during the initial 6-7 years of follow-up (Fig. 4B).

There are similarities between the present results and our recent analysis of the natural history of patients diagnosed with spinocerebellar ataxia type 3 (SCA3, or Machado-Joseph disease): LA/CA but not other antiparkinsonian drugs were associated with delayed SCA3 progression at 6 years and (to a less extent) 13 years of disease monitoring, in an effect that was adjusted for age and genetic burden, and that is possibly related to dopamine-mediated remodelling of protein aggregation pathways.^31^ It has been reported that adverse effects associated with long-term use of levodopa start to manifest in PD 6-7 years after the onset of Parkinson’s symptoms.^32^ Therefore, LA/CA treatments may benefit SCA3 and AD patients already showing aggravating signs of the disease, in line with the apparent delay in cognitive decline achieved for MCI subjects. Such a therapeutic strategy would also balance the increased risk of death that was identified in LA/CA-treated NC patients (Fig. 5A). Even if Parkinsonian symptoms are not manifested by most SCA3 or AD patients, the dopaminergic system is known to be affected in both diseases. ^2–5,33,34^ Thus, levodopa-based drugs with an adjusted dose to treat mild dopamine deficiency are conceived as possible approaches to delay the progress of SCA3 and AD in the future.

The caveats of the present study are connected with its observational and exploratory nature, lack of control of drug dosing and adherence, and limited control for comorbidities, concomitant medication and demographic variables.

Underreporting of treatments and/or mortality is an inherent risk arising from the self-reporting of medication use and the contingency that NACC is not always made aware of deaths for active and inactive subjects.^10,18,35^ Since the primary focus of the NACC-UDS is AD,^36^ Parkinsonian symptoms might be underreported even for subjects exposed to dopaminergic drugs. Another limitation was the lack of drug exposure data prior to entry into the NACC-UDS database or close to the dates of CSF biomarker analysis. The latter uncertainty is mitigated through the exclusion of biomarker data obtained more than 2 years after the reports of prescribed medication. Since LA/CA exposition was dichotomized between users and non-users, no insights could be provided into the effect of dosing on disease outcomes.

Conclusions about the best LA/CA strategies were also limited by the fact that patients prescribed with levodopa alone or with adjuntive therapies such as carbidopa, entacapone or dopamine agonists are all classified as LA/CA users. As signalled in previous retrospective studies,^15^ the nature of the NACC cohort and possible selection bias toward highly educated White participants may limit the general applicability of our findings. Therefore, the causality relationships hereby suggested between LA/CA use and disease outcomes require better-controlled clinical studies to be conclusively demonstrated towards the goal of effective and safe therapies for AD.

## Data Availability

All data produced in the present study are available upon reasonable request to the authors.

## Acknowledgements

This work was funded by National Funds through FCT—Fundação para a Ciência e a Tecnologia, I.P., under the project UIDB/04293/2020 and PTDC/QUICOL/2444/2021. PMM is supported by FCT CEECIND/03750/2017/CP1386/CT0014 (https://doi.org/10.54499/CEECIND/03750/2017/CP1386/CT0014).

The NACC database is funded by NIA/NIH Grant U24 AG072122. NACC data are contributed by the NIA-funded ADRCs: P30 AG062429 (PI James Brewer, MD, PhD), P30 AG066468 (PI Oscar Lopez, MD), P30 AG062421 (PI Bradley Hyman, MD, PhD), P30 AG066509 (PI Thomas Grabowski, MD), P30 AG066514 (PI Mary Sano, PhD), P30 AG066530 (PI Helena Chui, MD), P30 AG066507 (PI Marilyn Albert, PhD), P30 AG066444 (PI David Holtzman, MD), P30 AG066518 (PI Lisa Silbert, MD, MCR), P30 AG066512 (PI Thomas Wisniewski, MD), P30 AG066462 (PI Scott Small, MD), P30 AG072979 (PI David Wolk, MD), P30 AG072972 (PI Charles DeCarli, MD), P30 AG072976 (PI Andrew Saykin, PsyD), P30 AG072975 (PI Julie A. Schneider, MD, MS), P30 AG072978 (PI Ann McKee, MD), P30 AG072977 (PI Robert Vassar, PhD), P30 AG066519 (PI Frank LaFerla, PhD), P30 AG062677 (PI Ronald Petersen, MD, PhD), P30 AG079280 (PI Jessica Langbaum, PhD), P30 AG062422 (PI Gil Rabinovici, MD), P30 AG066511 (PI Allan Levey, MD, PhD), P30 AG072946 (PI Linda Van Eldik, PhD), P30 AG062715 (PI Sanjay Asthana, MD, FRCP), P30 AG072973 (PI Russell Swerdlow, MD), P30 AG066506 (PI Glenn Smith, PhD, ABPP), P30 AG066508 (PI Stephen Strittmatter, MD, PhD), P30 AG066515 (PI Victor Henderson, MD, MS), P30 AG072947 (PI Suzanne Craft, PhD), P30 AG072931 (PI Henry Paulson, MD, PhD), P30 AG066546 (PI Sudha Seshadri, MD), P30 AG086401 (PI Erik Roberson, MD, PhD), P30 AG086404 (PI Gary Rosenberg, MD), P20 AG068082 (PI Angela Jefferson, PhD), P30 AG072958 (PI Heather Whitson, MD), P30 AG072959 (PI James Leverenz, MD).

